# SIR model for assessing the impact of the advent of Omicron and mitigating measures on infection pressure and hospitalization needs

**DOI:** 10.1101/2021.12.25.21268394

**Authors:** Jan-Diederik van Wees, Martijn van der Kuip, Sander Osinga, Bart Keijser, David van Westerloo, Maurice Hanegraaf, Maarten Pluymaekers, Olwijn Leeuwenburgh, Logan Brunner, Marceline Tutu van Furth

## Abstract

**Background:** On 26 November 2021, the world health organization (WHO) designated the coronavirus SARS-CoV-2 B.1.1.529 a variant of concern, named Omicron (WHO, 2021a). As of December 16, Omicron has been detected in 89 countries (WHO, 2021b). The thread posed by Omicron is highly uncertain.

**Methods and findings:** For the analysis of the impact of Omicron on infection pressure and hospitalization needs we developed an open-source stochastic SIR (Susceptible-Infectious-Removed) fast-model for simulating the transmission in the transition stage from the prevailing variant (most often Delta) to Omicron. The model is capable to predict trajectories of infection pressure and hospitalization needs, considering (a) uncertainties for the (Omicron) parametrization, (b) pre-existing vaccination and/or partial immunity status of the population, and demographic specific aspects regarding reference hospitalization needs, (c) effects of mitigating measures including social distancing and accelerated vaccination (booster) campaigns.

**Conclusions:** The SIR model approach yields results in fair agreement with Omicron transmission characteristics observed in South Africa and prognosis results in Europe (UK and Netherlands). The equations underlying the SIR formulation allows to effectively explore the effect of Omicron parametrization on anticipated infection growth rates and hospitalization rates relative to the prevailing variant. The models are online available as open source on GitHub.

**One Sentence Summary:** fast-model for the impact of Omicron

## Introduction

On 26 November 2021, the world health organization (WHO) designated the coronavirus SARS-CoV-2 B.1.1.529, a variant of concern, named Omicron (WHO, 2021a). As of December 16, Omicron has been detected in 89 countries (WHO, 2021b). The thread posed by Omicron is highly uncertain. According to the World Health Organization the overall threat posed by Omicron largely depends on four key questions, including: (1) how transmissible the variant is; (2) how well vaccines and prior infection protect against infection, transmission, clinical disease and mortality; (3) how virulent the variant is compared to other variants; and (4) how populations understand these dynamics, perceive risk and follow control measures, including public health and social measures.

Irrespective of these critical uncertainties, governments and health organizations are required to timely implement measures that are both effective, and least detrimental for society and economy. It is thus of importance that public health advice can be based on most recent insights in characteristic of the variant of concern and is tailored according population-specific characteristics. In this study, we established an open-source epidemiological model allowing to predict infection and hospitalization rates inferring parametrized viral transmission.

The rationale behind this paper is to provide a simple yet effective and transparent predictive tool, based on a SIR (Susceptible-Infectious-Removed) model formulation, which can easily be used to anticipate the potential effects of the transition to Omicron in terms of prognosed infections and hospitalization needs. Furthermore, it allows to analyze the effects of social distancing and accelerated booster campaigns to mitigate adverse effects. Transparency and ease of access to the model is provided through (a) detailed description of the used SIR model, (b) open source Python distribution (https://github.com/TNO/TNO-COVID-Variant-SIR/), (c) analysis of common model-derived characteristics of Omicron and their effect on Omicron’s growth rate and impact, and (d) illustration of its value in exemplary (synthetic) case studies.

The paper is structured as follows: First, we introduce the model assumptions, and common characteristics and parametrization, and their bearings on Omicron transmission characteristics and case hospitalization rates. Next, we illustrate the model for a number of (synthetic) case studies in South Africa (Gauteng State), the UK and the Netherlands. These settings are considered representative for markedly different demographic conditions, vaccine status, natural immunity and capability for booster campaigns. The prime intention of these studies is to demonstrate the proper functioning of the model, and its capability to predict first order infection and hospitalization trends in line with data and other models.

## Model assumptions

We adopt an epidemiological compartmental Susceptible-Infected-Removed (SIR) model which is as well as the closely related SEIR (where the E stands for exposed) model, a classic model for transmission which has been used extensively for COVID-19 model predictions (e.g. Van Wees et al., 2020, Cooper et al., 2020; Yang and Shaman, 2021). The SIR model is detailed in the supplemental materials, and takes into account the effects of existing (double) vaccination, enhanced protection with boosters, and a simple mathematical formulation for the transition from the prevailing variant (Delta in most countries) to Omicron.

For the transmission model we adopt similar parametrization conventions and parameter value ranges as proposed in Barnard et al. (2021). We compiled a range of vaccine and booster efficacies, and other parameters relevant for immune evasion and case hospitalization rates from recent literature and reports in table 1. More general parameter ranges of relevance for the model (case studies) are shown in table 2.

**Table 1.** parameters for the model first listed parameter. ^1^values for South Africa (SA) correspond to relative recent vaccinated (10-14 weeks Pfizer in Figure 10 of UKHSA,2021c); for the Netherlands (NL), which is representative for vaccination status for continental Europe, is marked by relatively waned vaccines (25+ weeks Pfizer in Figure 10 of UKHSA,2021c) and very recent Pfizer boosters (cf Figure Figure 10 of UKHSA,2021c) UK is marked. ^2^ Studies on (waning) vaccine and booster efficacy, ^3^data from the Netherlands. ^4^50% hospitalization compared to Delta (UKHSA 2021c, ISS,2021), in SA ca 40% based on data fit (Figure 3). ^5^Barnard et al. (2021). ^6^value following Yang and Shaman, 2021b, and in close correspondence to vaccine efficacy for infection.

| Variant | Parameter | Value SA | Value UK | Value NL | Symbol | Json parameter |
| --- | --- | --- | --- | --- | --- | --- |
| Delta | Vaccine efficacy infection | 0.75<sup>1</sup> | 0.75<sup>1</sup> | 0.6<sup>1</sup> | $V_{inf_d}$ | ve_vac_d |
| Omicron | Vaccine efficacy infection | 0.25<sup>1</sup> | 0.25<sup>1</sup> | 0.1<sup>1</sup> | $V_{inf_o}$ | ve_vac_o |
| Delta | Booster efficacy infection | na | 0.9<sup>1</sup> | 0.9<sup>1</sup> | $B_{inf_d}$ | ve_booster_d |
| Omicron | Booster efficacy infection | na | 0.7<sup>1</sup> | 0.7<sup>1</sup> | $B_{inf_o}$ | ve_booster_o |
| Delta | Vaccine efficacy hospitalization | 0.95<sup>2</sup> | 0.9<sup>3</sup> | 0.9<sup>3</sup> | $V_{hosp_d}$ | ve_vac_hosp_d |
| Omicron | Vaccine efficacy hospitalization | 0.975<sup>7</sup> | 0.9-0.95<sup>4</sup> | 0.9-0.95<sup>4</sup> | $V_{hosp_o}$ | ve_vac_hosp_o |
| Delta | Booster efficacy hospitalization | na | 0.95<sup>2</sup> | 0.95<sup>2</sup> | $B_{hosp_d}$ | ve_booster_hosp_d |
| Omicron | Booster efficacy hospitalization | na | 0.95-0.98<sup>3</sup> | 0.95-0.98<sup>3</sup> | $B_{hosp_o}$ | ve_booster_hosp_o |
| Omicron | Relative protection of re-infected against hospitalization compared to delta (1= complete protection) | 0.6<sup>4</sup> | 0.3-0.7<sup>4</sup> | 0.3-0.7<sup>4</sup> | $U_{hosp_o}$ | ve_immune_hosp_o |
| | Vaccinated fraction | 0.3 | 0.55 | 0.8 | $p_v(0)$ | p_vac_start |
| | Boostered fraction (at starting time of simulation minus $t_{shift}$) | na | 0.2 | 0.0 | $p_b(0)$ | p_booster_start |
| Delta/Omicron | Vaccine/booster efficacy transmission | 0.4<sup>5</sup> | | | $V_{trans}$ | ve_trans |
| Omicron | Immune protection for passed infections | 0.33<sup>6</sup> | | | $U_{inf_o}$ | ve_immune_o |

**Table 2.**
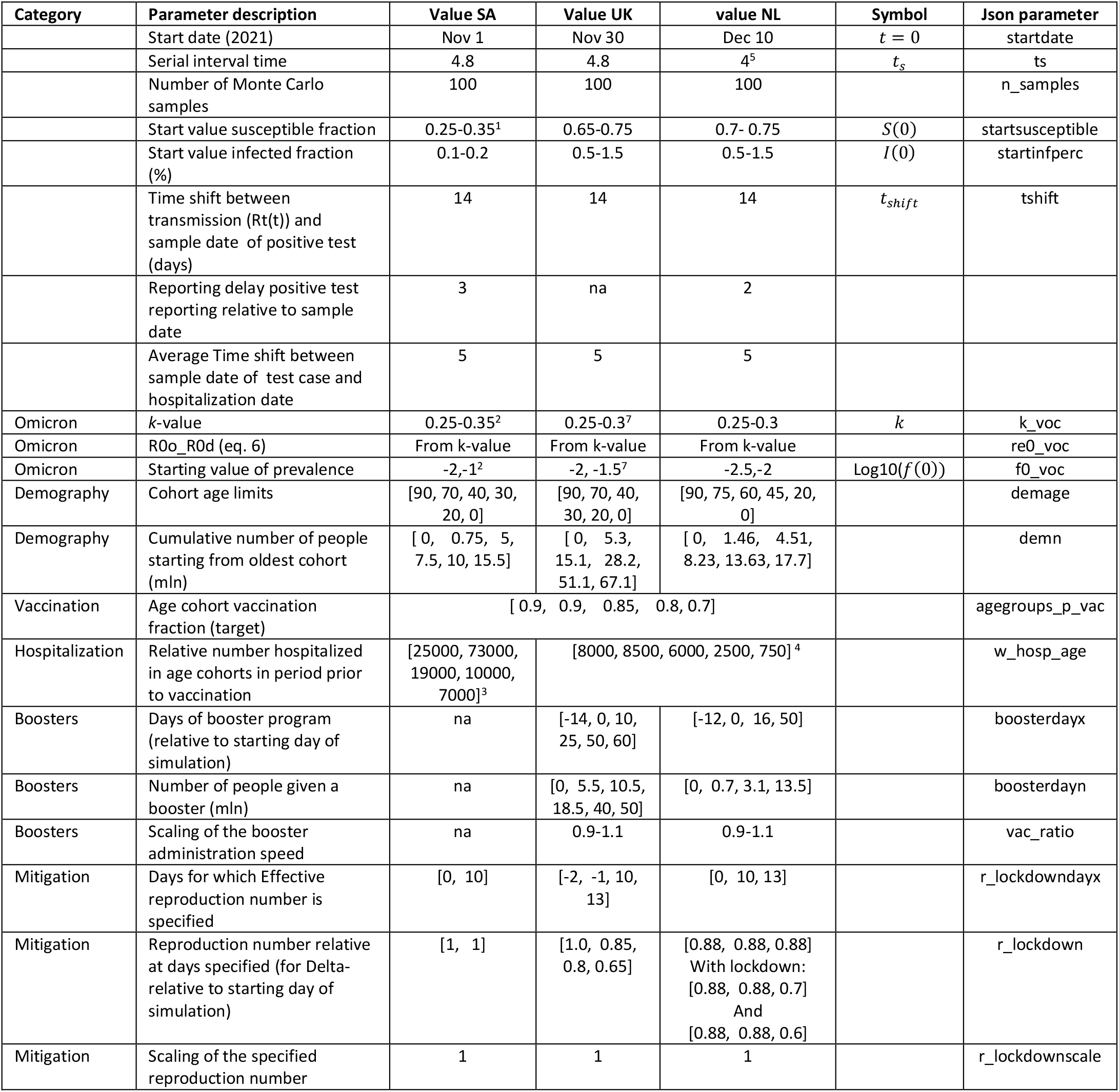
parameters for the model. ^1^in accordance with Yang and Shaman, 2021b, ^2^following Pearson et al., 2021. ^3^from NIDC (2021) cumulative data estimates.^4^from NICE (2021), data 6 Januari 2021. ^5^serial interval time used for reporting by RIVM (2021a). ^7^ based on data from GOVUK, 2021 (see Fig. 4)

An alarming feature of Omicron is the extremely fast growth rate of the prevalence of Omicron to Delta doubling each 2-3 days in the UK (UKHSA,2021a,2021b), Denmark (ISS,2021), Belgium and the Netherlands (RIVM, 2021c).

According to WHO (2021b), Omicron is spreading rapidly in countries with high levels of population immunity (either through natural infection or vaccine protection) and it is becoming more and more clear that the rapid growth rate should be primarily attributed to immune evasion. Barnard et al. (2021), based on transmission model considerations in accordance with observed growth of prevalence of Omicron to Delta, arrived at a Basic Reproduction number for Omicron which is no more than 30% higher compared to Delta. Such a low ratio demonstrates immune evasion as the dominant mechanism for transmission, in order to explain the observed growth rates. Using Eq.6 from the supplement, we used the listed parameter values in Table 1 to determine expected growth rates of Omicron relative to Delta (Figure 1) as a function of the current vaccination status, adopting a 20% higher reproduction number for Delta. The figure illustrates very well the clear difference in characteristics of growth as function of relative immunity loss in a setting largely determined by passed natural infections (SA) *vs* dominated by vaccinations (NL). In SA the growth is mainly caused by immunity loss from passed infections. In the EU (e.g. Netherlands) this component is less significant due to the vaccinations, and strongest relative growth of omicron is related to immunity loss from a high concentration of boosters and from passed infections. The latter effect could possibly explain very high growth rates in cities with young population as observed in the United Kingdom in London with k∼0.4 (UKHSA, 2021b) and the onset of Omicron in Denmark (ISS, 2021).

**Figure 1.**
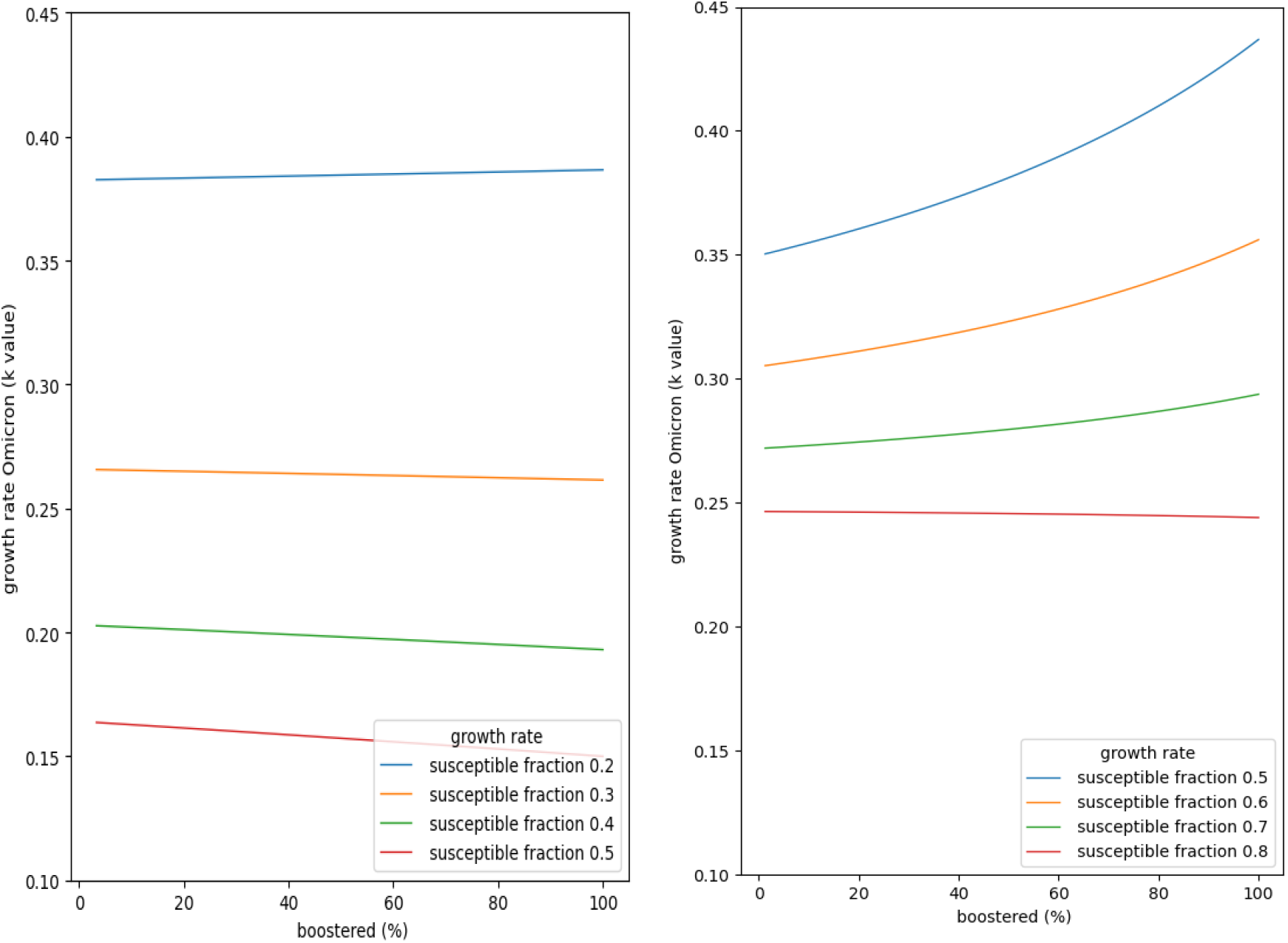
(left) expected growth rate for Omicron in countries with immunity from past infections, through past waves, adopting median parameter values listed for South Africa in Table 1, and 20% increase of reproduction number of Omicron relative to Delta. (right) for mean parameter values listed for the Netherlands in Table 1. The susceptible fraction is varied to show the dominant sensitivity to immunity loss of past infections. The horizontal axis is scaled to the booster fraction relative to the vaccinated fraction. So, 100% means all vaccinated received a booster.

In South African province Gauteng, infection-rates (positive tests) increased almost 100 fold from early November, peaking early December (https://ibz-shiny.ethz.ch/covid-19-re-international/), whereas the weekly reported hospitalization rates in the corresponding weeks peaked only at a 20 fold increase in week 49 (NICD, 2021). Recent data analysis of Omicron cases in the UK, also seems to indicate an significant reduction in case hospitalization rates of approximately 50% relative to Delta (Ferguson et al., 2021a; Ferguson et al., 2021b, UKHSA, 2021c). Similarly, also most recent data in Denmark indicates that hospitalization rates are lower compared to Delta. Still there is uncertainty since the numbers of Omicron cases are relatively small, and lab analysis is not conclusive on a possible reduced protection of vaccines against severe illness.

## Exemplary case studies

The practical functioning of the SIR model is demonstrated in two exemplary case studies: in South Africa (Gauteng province; estimated population 15,8 million; area 18,000 km^2^), the UK (estimated population 67.1 mln; area 242,500 km^2^) and the Netherlands (estimated population 17,7 million, area 42,000 km^2^). The parametrization for both model simulations is given in Tables 1 and 2.

The model performs a number of Monte-Carlo runs, allowing to visualize and analyze the bandwidth of predictions as a function of uncertainty in the listed parameters (all indicated ranges are treated as uniformly distributed).

The model takes into account the demographic aspects through the specification of age cohorts, the specification of the vaccinated fractions in these age cohorts, and their hospitalization needs in the non-vaccinated reference situations (prior to the advent of vaccines). The predicted infections and hospitalizations are in this version of the model not convolved with a gamma function which is required to realistically mimic delay and variability in timing of development of symptoms into illness and hospitalization (e.g. Van Wees et al., 2020). Furthermore, the current model neglects time delays in registration. These would result in a shift of approximately a week for positive cases and two weeks for hospitalization and would result in significantly longer tails than shown in the figures.

The timing and strength of Omicron is determined by the input *f0* and *k*-value respectively. Using equation 6 the ratio of the (basic) reproduction number of Omicron and Delta is calculated and subsequently used in the SIR model simulation characteristics.

For mitigating measures two options can be implemented. Firstly, social distancing measures can be specified which are interpreted to result in a reduction of the reproduction number as if Omicron would be absent (i.e. in the Delta reference). Secondly, boosters can be administered.

### Prediction of infection and hospitalization rates in the South African province Gauteng

In South Africa, the Omicron variant was identified in the last weeks of November, and the variant originated at least prior to the start of November. The population of South Africa is marked by a relatively young age and only approximately 30% of the population has been vaccinated (We assume elderly people have been assumed with a vaccination rate of ca 90%). The Delta wave has caused most people to have been exposed to the virus and have buildup natural immunity to delta (estimated 60-65%), taking into account immune evasion of past waves (Yang and Shaman, 2021a, 2021b). The starting effective reproduction number has been set to 1.0, and possible time variant effects of social distancing measures and any possible effect of growing vaccination has been ignored.

**Figure 2.**
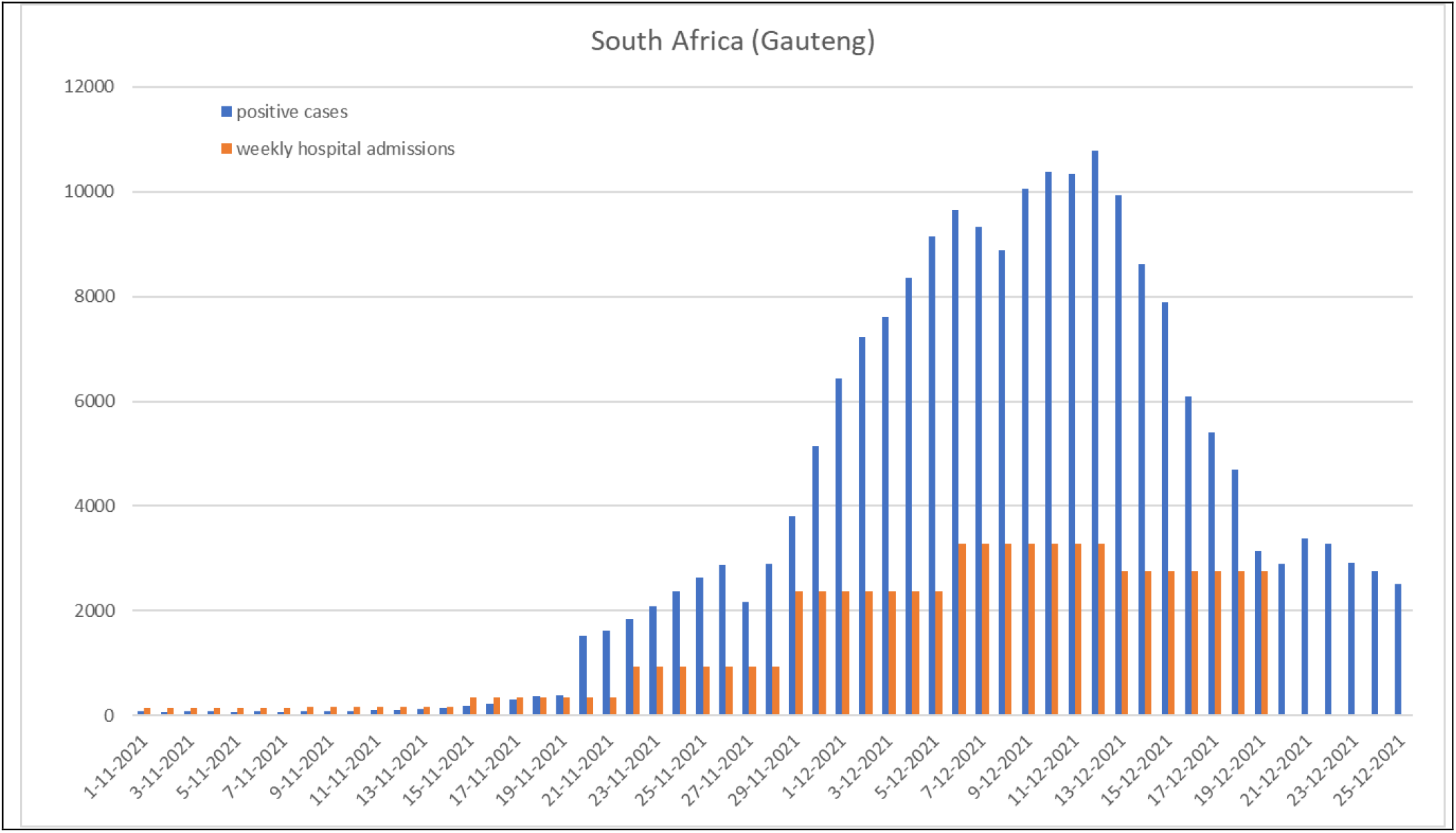
evolution of (weekly averaged) registered cases and weekly hospital admissions in Gauteng due to Omicron becoming dominant in second half of November, marked by a sharp increase in positive cases. Data sources https://dsfsi.github.io/covid19za-dash/ and NIDC(2021)

The results of the simulation predict both the 100 fold increase in cases as an effect of immune evasion, and the observed hospitalization rates which are marked by a 20 fold increase in the first weeks of December compared to the early November.

**Figure 3.**
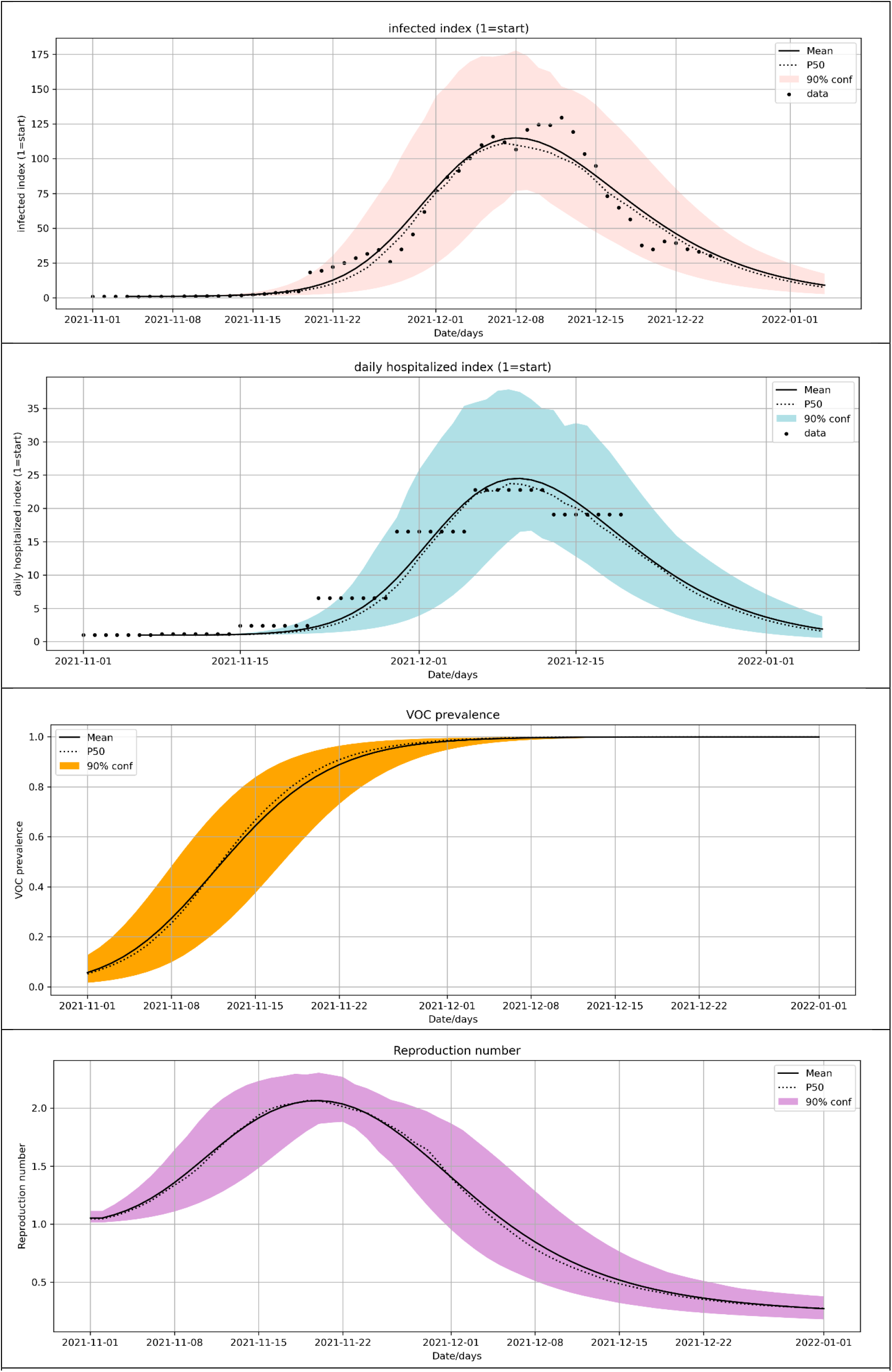
model of Gauteng. (top) relative growth of infections and (second from top) translation to corresponding hospitalization rates (neglecting temporal convolution of infection rates to hospitalization. (third from top) Omicron prevalence. (bottom) effective reproduction number. Please note that for the latter curve a timeshift should be applied of -t_shift_=-14 days

### Prediction of infection and hospitalization rates in the UK

In the UK Omicron has been marked by a rapid growth of Omicron (UKHSA, 2021a; Barnard et al., 2021), being dominant from the second half of December 201 onwards (GOVUK, 2021; Figure 4). The population has been for over 75% fully vaccinated and almost half of the fully vaccinated received a booster. Consequently the UK population was rather well protected against the Delta variant and until the end of November the infection and hospitalization rates have been rather stable.

**Figure 4.**
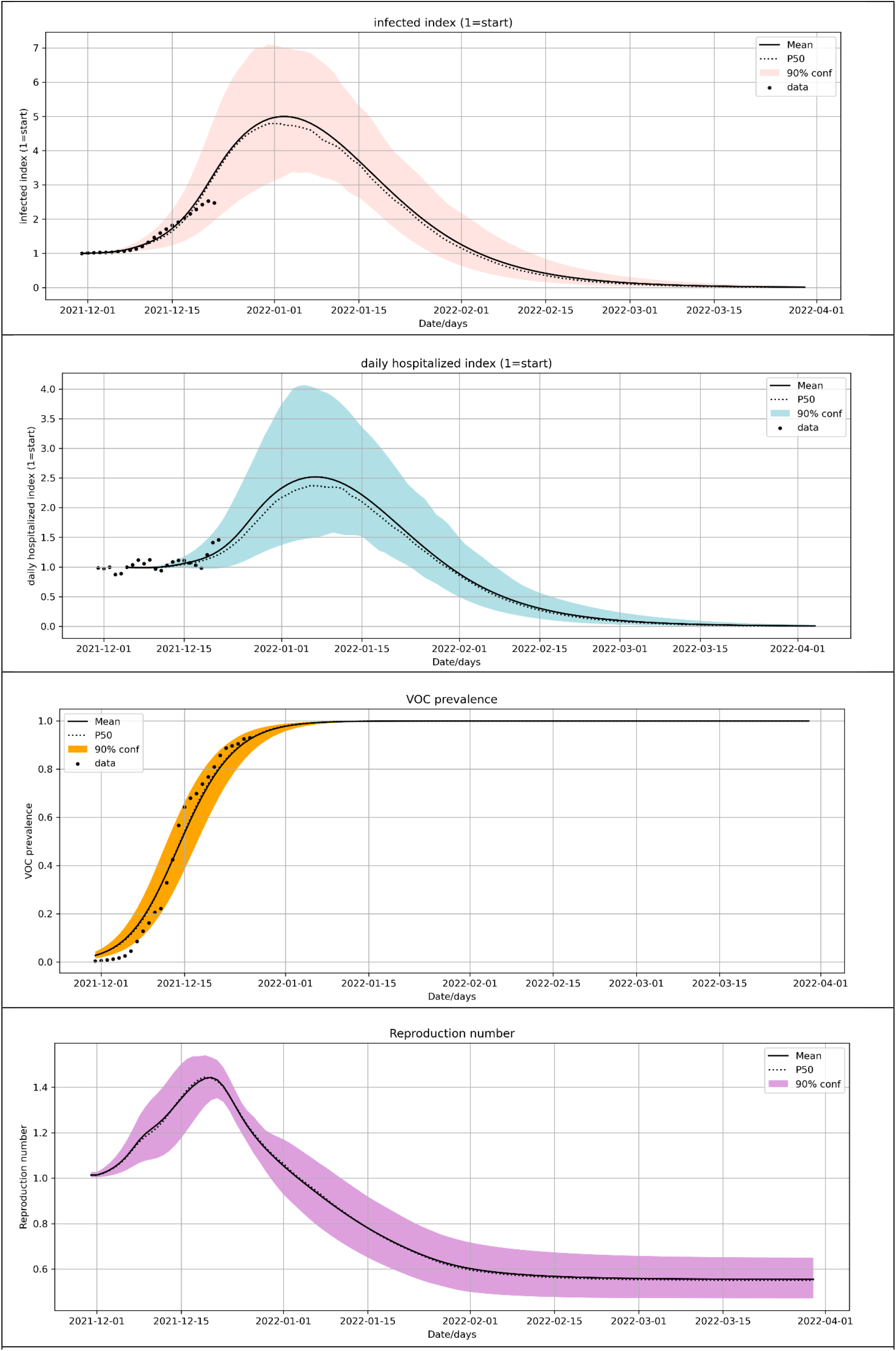
model of UK. (top) relative growth of infections (plotted with weekly average from data) and (second from top) translation to corresponding hospitalization rates (with data not weekly averaged). (third from top) Omicron prevalence. (bottom) effective reproduction number. Please note that for the latter curve a timeshift should be applied of -t_shift_=-14 days. Data are according to registration date from GOVUK (2021)

With the advent of Omicron, the UK government re-instated from end of November a number of measures including wearing facemasks in public transport to reduce the transmission, and also to accelerate the ongoing booster campaign to improve protection.

From 10 December onwards, intensified -so called Plan B-measurements were taken, including wearing face coverings in public buildings, and advice to work from home.

The number of infections kept stable for another week, but started to rise the last two weeks, and more recently also the hospitalization rates.

In the model (Figure 4) the effect of social distancing measures are picking up about Christmas, and are likely to have a positive but yet unknown effect.

The chosen k-value is systematically lower than observed in individual growth curves reported at low prevalence rates below 10% (UKHSA, 2021b). This may be attributed to geographic spreading effects as well a possible effects of change in transmission from predominantly higher susceptible groups to higher susceptible groups (Fig. 1, right panel)

### Prediction of infection and hospitalization rates in the Netherlands

In the Netherlands, the hospitalization rates just have reached a peak in early December and are declining, at the same time Omicron is marked by community spreading at relatively large rates, with reported prevalence of approximately 1% nationally reported at December 10 (RIVM, 2021b). In the Netherlands, about 80% of the population is fully vaccinated, the majority with two doses of Pfizer. A booster campaign has started end of November with 2% of the population boostered early December. In response to Omicron the campaign has recently been accelerated with plans to administer boosters to all vaccinated adults (18+) at the end of January 2022. In the model this has been included.

The initial effective reproduction number is R=0.88 (RIVM, 2021c). The prognosed infection rates and projected hospitalization needs (Figure 5) are marked by a strong anticipated growth, due to the large uncertainties of the characteristics of the Omicron variant. However, with 90% confidence interval the hospitalization rates for the R=0.88 scenario would grow considerably higher than the rates observed today, despite the accelerated booster campaign. Therefore the Dutch government decided to impose a strong lock down effective from 19 December onward with a significant anticipated reduction of the effective reproduction number.

**Figure 5.**
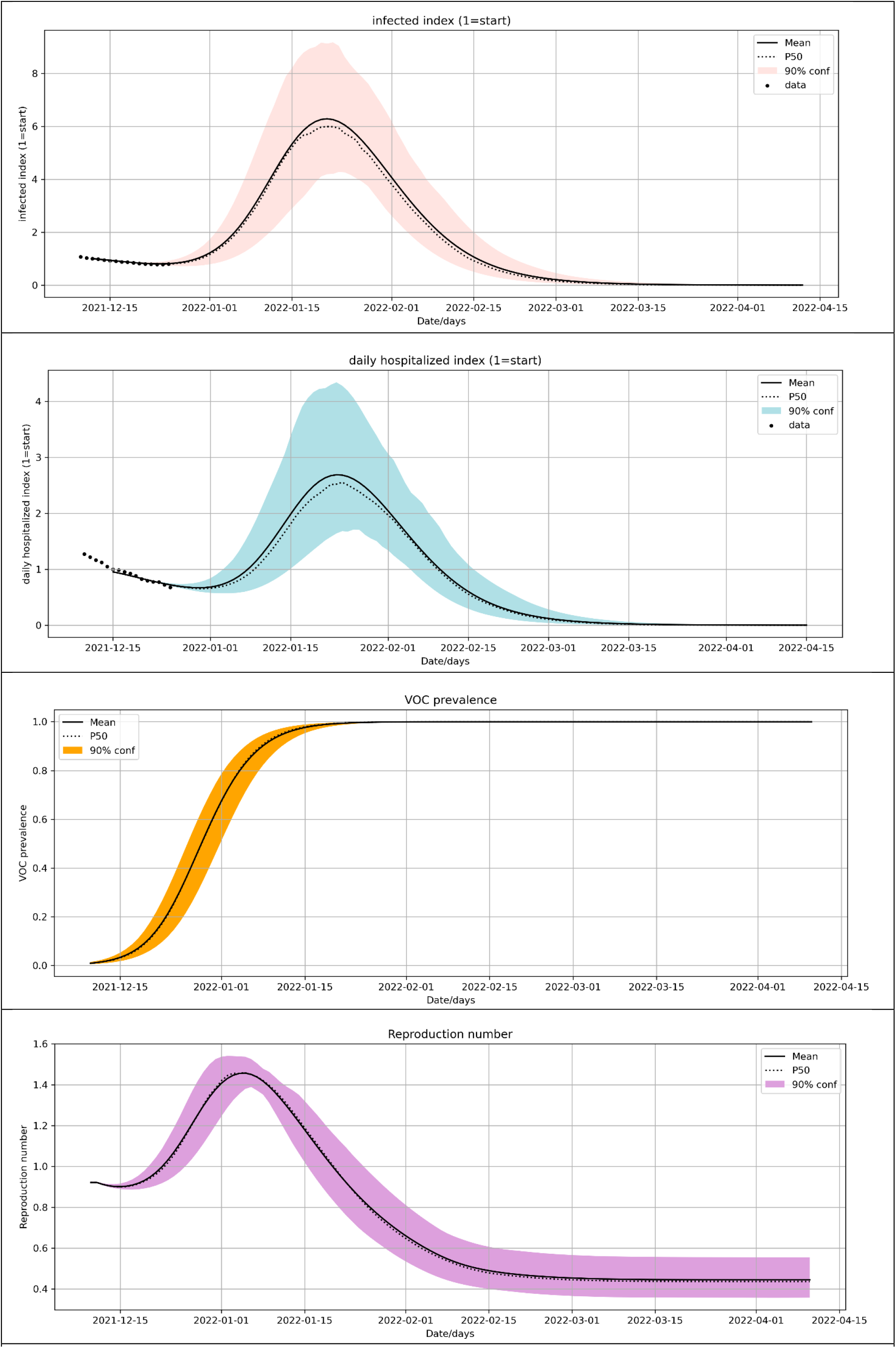
model of the Netherlands, for R (delta)=0.88 (same conventions as in Figure 3, data are weekly averaged, positive cases are according to reporting date, hospitalization registration date)

The updated scenario for a lockdown with R=0.6 and R=0.7 accomplished in the 5 days following December 19 (Figure 6).

**Figure 6.**
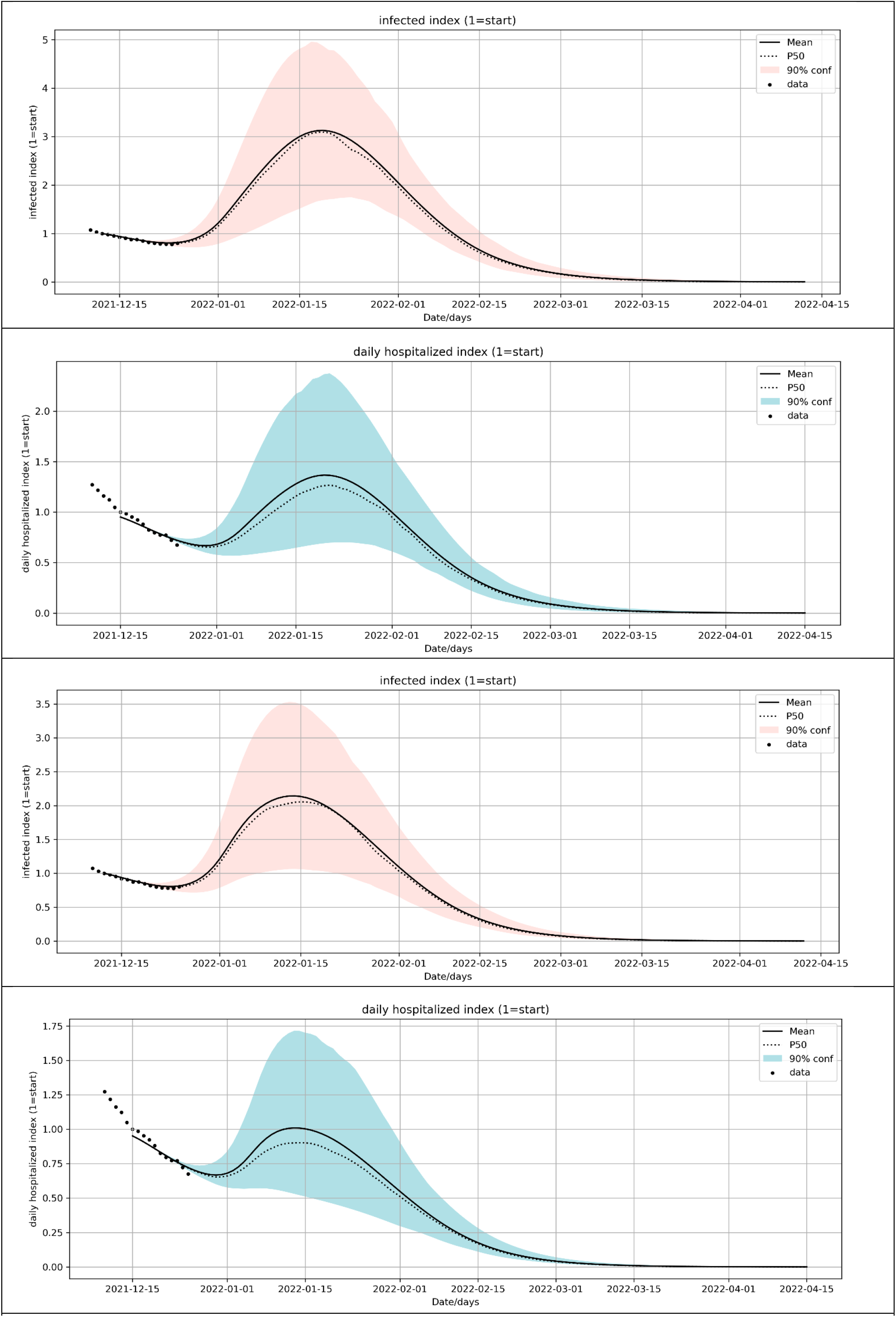
model of the Netherlands, for lockdown R=0.7 (top panels) and R=0.6 (bottom panels)

In order to understand the role of different uncertainties and their control on the potential peak in hospitalization needs we did run 10,000 Monte-Carlo realizations and generated cross plot for the varying parameters and their effect on the peak hospitalization needs. The results have been shown in Figure 7. The sensitivity analysis shows that the uncertainty in the peak hospitalization needs are strongly correlated with *k*-value (k_voc) and the interpretation of start infection rate (startinfperc), results in very large uncertainty. In addition, many other parameters correlate with peak hospitalization including the achieved speed in booster administration (vac_ratio), the starting prevalence of Omicron (f0).

**Figure 7.**
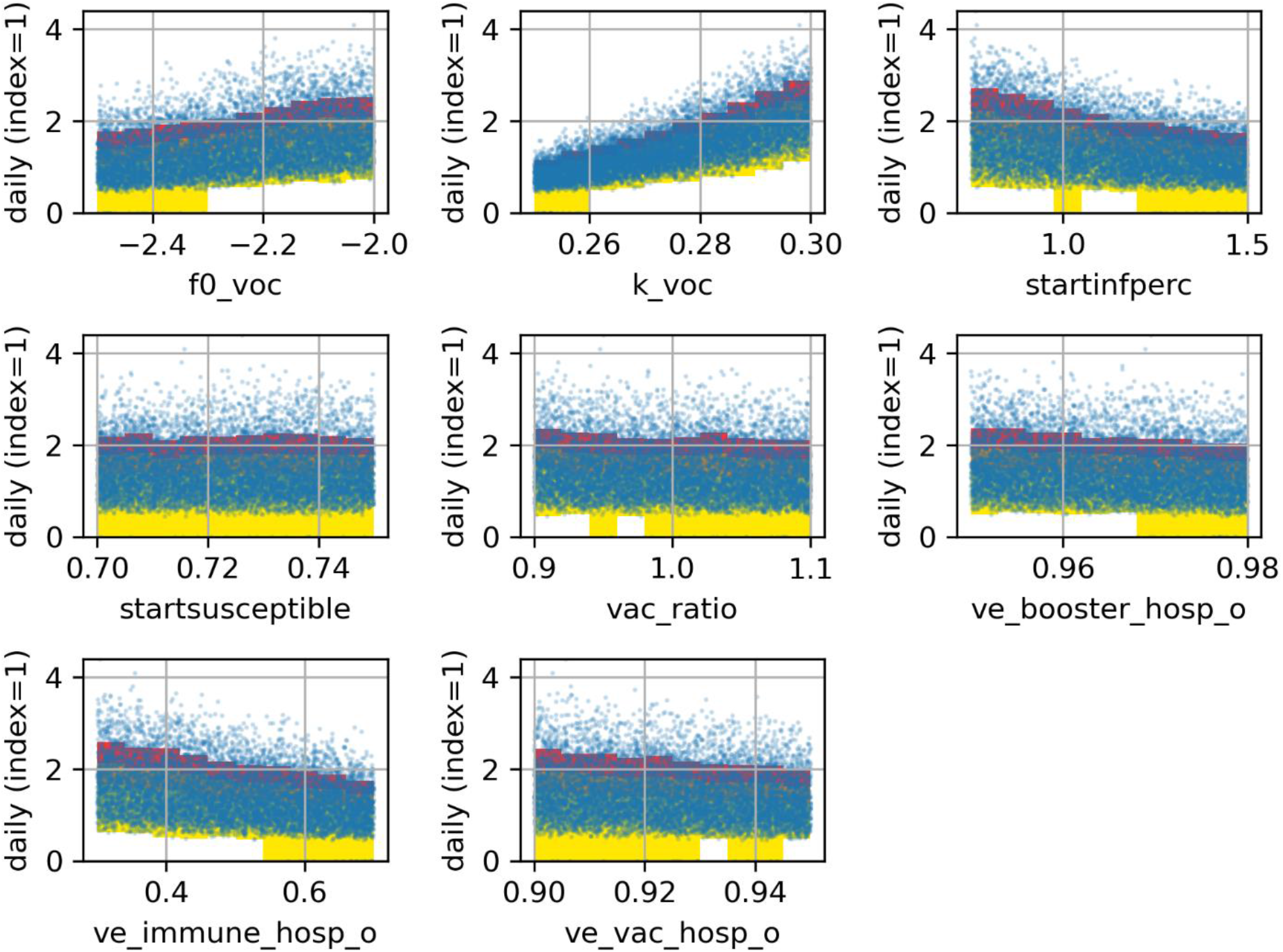
sensitivity analysis for peak hospitalization predictions for the Netherlands for R=0.7 lockdown scenario. Dots correspond to individual outcomes. Colors correspond to threshold levels of 25% (red, upper bound 10%), 50%(orange), and remainder (yellow).The variable names correspond to the names in json column of tables 1 and 2.

The models demonstrate very clearly that without additional measures the Omicron wave would pose a major threat to the health care system. This is perfect agreement with results from more sophisticated transmission models of the Dutch Health agency (RIVM, 2021b), which at the time publication did not yet include the effects adjustment towards lower case hospitalization rates. These findings also concur with model studies in the UK, which concluded the need for more stringent social distancing measures,as being pursued with Plan B (Barnard et al., 2021).

## Conclusions

We presented in this paper a simple yet effective and transparent predictive tool, which can easily be used to anticipate the potential effects of the transition to Omicron in terms of prognosed infections and hospitalization needs. Furthermore, it allows to analyze the effects of social distancing and accelerated booster campaigns to mitigate adverse effects.

We presented an analysis of common model-derived characteristics of Omicron and their effect on Omicron’s growth rate and impact and illustrated its value in exemplary (synthetic) case studies.

The model and the shown example case studies are available as open source Python distribution on GitHub (https://github.com/TNO/TNO-COVID-Variant-SIR/)

## Data Availability

All data produced are available online at the data availability link

https://github.com/TNO/TNO-COVID-Variant-SIR/

## Supplement

### SIR model for the transition-stage from delta to omicron, including the effects of a booster campaign

For the transmission model to predict the changes of infection rate over time *t* in days, we integrate over time an epidemiological compartmental model ODE formulation (*t=0* corresponds to the start of the simulation time):

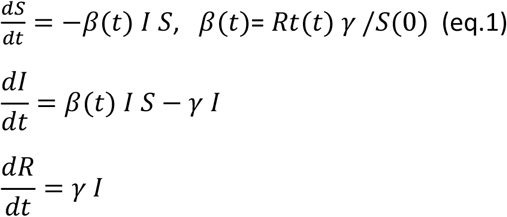

*S(t), I (t), R(t)* are susceptible, infected and removed compartments (fractions) of the population, *γ* = 1/*t*_*s*_. *S+I+R* =1. *R(0*) is an estimate of the fraction of the population which has been infected and recovered from delta at the start of the simulation The fraction of population which have died is ignored. *I (0)* is estimated from the current infection rate multiplied by *t*_*s*_.

The *Rt*(*t*) is the effective reproduction number taking into account the prevalence of the Variant of Concern *f* (*t*), marked by subscript o for Omicron, whereas the prevalent variant is denoted *d* for delta

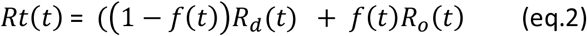

*f* (*t*) is growing prevalence of omicron:

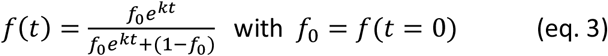

Where *k* is daily growth rate of the natural logarithm of *f* (*t*), *f*_0_ is fraction of omicron at starting time. *k* can be determined from observations of growth rate of omicron

For the prevailing delta variant we can write the expected reproduction number as *R*_*d*_ (*t*) in response to ongoing vaccination, which is the compound effect of the expected change in infection rate *I*_*d*_(*t*), and the expected change in transmission rate *C*_*d*_(*t*) :

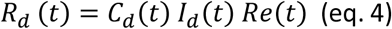

Where

- If we assume the population to be perfectly mixed (this is used in the calculation)
  - 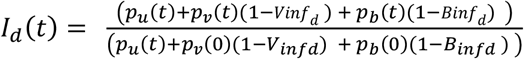
  - 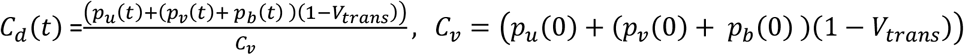
  - Or alternatively segregated
  - 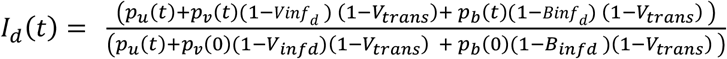
  - *C*_*d*_(*t*) =1
- *p*_*u*_(*t*), *p*_*v*_(*t*), *p*_*b*_(*t*) are unvaccinated, vaccinated, and boostered fractions of the population
- *Vinf*_*d*_ is the average vaccine efficacy (of the (already) double vaccinated fraction of the population) for the delta and omicron variant. *Binf*_*d*_ is the vaccine efficacy of newly administered (and existing) boosters against infection for the delta.
- *V*_*trans*_ is the vaccine efficacy for transmission and is assumed equal for both re-infected, vaccinated and boostered people (cf Barnard et al., 2021).

For Omicron we can write:

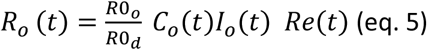

Where

- 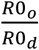 is the ratio of the basic reproduction number of omicron and delta. This ratio can be determined form the growth rate of omicron prevalence
- If we assume the population to be perfectly mixed (this is used in the calculation)
  ◯ 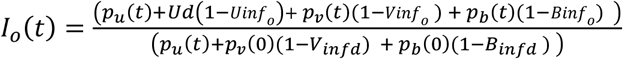
  ◯ 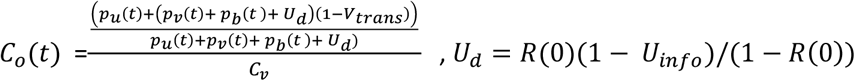 Alternatively for segregated
  ◯ 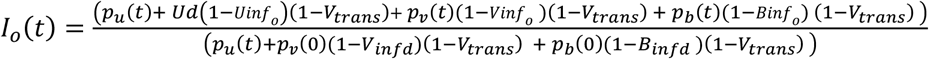
  ◯ *C*_*o*_(*t*) =1
- *Vinf*_*o*_ and *Binf*_*o*_ is the vaccine efficacy of newly administered (and existing) boosters respectively against infection for the omicron variant.
- *Uinf*_*o*_ is the protection against immunity loss for the recovered fraction for Omicron

We can write the apparent increase of the effective Reproduction number compared to the prevailing:

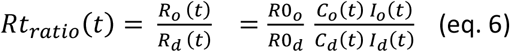

Now we can relate the observed growth rate *k* (the growth rate of the natural logarithm of *f(t)*) of omicron relative to delta to the *Rt*_*ratio*_(*t*) as follows from the SIR model characteristics normalized to S(0)=1, which holds

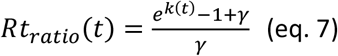

which follows from the effect on daily growth rate on the *β*-*γ* → *β* = *e*^*k*^ -1 + *γ*, and the related R value of the SIR model 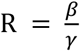

This means that the observed growth rate *k* at t=0 can be used to calibrate 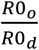 given other parameters in (eq. 7):

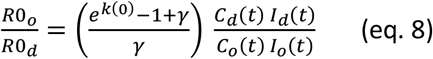

Reported growth rates vary from k=0.35 in the UK (UKHSA, 2021a) to lower numbers in NL (0.28), DK (0.30) and Belgium (0.28) (RIVM, 2021c) For a fixed 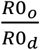, equation 8 demonstrates that observed growth rates are a function of the relative strength of immune evasion in the different compartments unvaccinated, vaccinated and boostered, and the ratio of recovered and susceptible fraction prior to the introduction of the variant.

### Hospitalization rates

The case hospitalization rates are dependent on different age groups, and differ for Delta and the Omicron variant

For each age group (*i*_*age*_) the case hospitalization rate *CHR*_*iage*_(*t*) is determined as follows

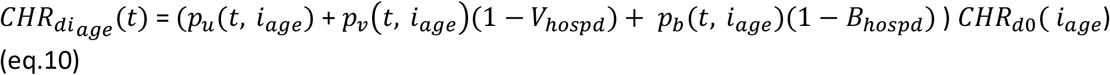

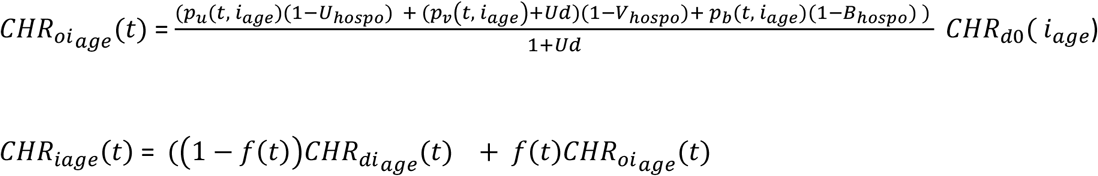

Where

- *p*_*u*_(*t, i*_*age*_), *p*_*v*_(*t, i*_*age*_), *p*_*b*_(*t, i*_*age*_) are relative fractions (summing to 1) of unvaccinated, vaccinated and boostered in the age group.
- *U*_*hospo*_ is the general reduction in hospitalization rate
- *V*_*hospd*_, *V*_*hospo*_ are vaccine (and reinfection) efficacies against hospitalization for delta and omicron
- *B*_*hospd*_, *B*_*hospo*_ are booster efficacies against hospitalization for delta and omicron
- *CHR*_*d*0_(*i*_*age*_) is reference hospitalization rates per age group for the delta situation

The total hospitalization rates becomes

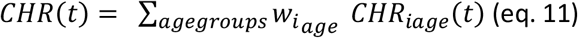

Where 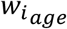 is the demographic fraction of the age group in the total population.

For the relative change in hospitalization rates it is not necessary to know the absolute *CHR*_*d*0_(*i*_*age*_) as its scaling drops out in the relative scaling (*CHR*(*t*) /*CHR*(0)),, so we can use arbitrary indicative numbers (i.e. hospital intakes from different age groups collected over the past waves periods)

### SIR model correction for realistic Incubation times, time delays in testing and registration

In the SIR model models the infection rate follows almost instantly the transmission rate defined through *Rt(t)*. However for prediction of positive cases, the model needs to take into account an appropriate time delay (or convolution) for delay time for transmission (the SIR approach is marked by a more direct transmission effect, compared to a SEIR formulation which is more realistic) and incubation time as well as delay in testing and reporting of positive cases. For the total delays we adopt on average *t*_*shift*_ =14 days. For case registration date we add an additional reporting delay. For development of severe illness leading to hospitalizations we assume on average 5 days on top of the 14 days.

In order to correct for these times we use the observed *f(t*) function (taking the dates from registered cases on which f(t) is based as reference model time). Any non-pharmaceutical measures or vaccination and their effect is therefore incorporated at time *t* = *t* + *t*_*shift*_ in the model, in order to take into account the correct time delay for transmission effects. For hospitalization the same argument is used, so hospitalization protection is in accordance with vaccine protection at the incubation stage.

COVID-19 Incubation times are lognormally distributed (i.e. Paul and Lorin, 2021 for an overview). In order to take these effects into account, we adopt a convolution of the predicted infections (and hospitalizations) with a lognormal function with a log mean value of 1.8 and standard deviation of 0.53. The mean value of the resulting lognormal distribution for the incubation time is 7 days. In order to be consistent with the *t*_*shift*_ approach, the convolution result is shifted -7 days. The convolution is also applied to the *1-f(t))* and *f(t*) and to construct the observed *f(t*) function in accordance with the expected convolution effects.

